# Effectiveness of a telenursing intervention program in reducing exacerbations in patients with chronic respiratory failure receiving noninvasive positive pressure ventilation: A randomized controlled trial

**DOI:** 10.1101/2022.05.30.22275763

**Authors:** Makoto Shimoyama, Fumiko Sato, Naoko Sato, Shiori Yoshida, Chikako Takahashi, Mizue Inoue

## Abstract

Telenursing for patients with chronic respiratory failure receiving noninvasive positive pressure ventilation (NPPV) is an important aid in reducing exacerbations; however, there is insufficient evidence. This randomized controlled trial aimed to investigate the effectiveness of a telenursing intervention program in reducing exacerbations in patients with chronic respiratory failure receiving NPPV at home. This study included patients who were receiving NPPV at home and could handle a tablet device. The intervention group (n = 15) was exposed to an information and communications technology-based telenursing intervention program, in addition to usual care; the control group (n = 16) received the usual care only. The telenursing intervention program comprised telemonitoring and health counseling sessions via videophone. The intervention was evaluated once at enrollment and after 3 months. The primary endpoints were the number of unscheduled outpatient visits, hospitalizations, and hospital days. The secondary endpoints included the St. George’s Respiratory Questionnaire (SGRQ), Euro QOL 5 Dimension, Self-Care Agency Questionnaire (SCAQ), pulmonary function tests, and 6-minute walking distance. We found no significant differences between the intervention and control groups at enrollment. At follow-up, the number of routine outpatient visits for acute exacerbations (r = .36; p = .045), the number of hospitalizations (r = .38; p = .037), the number of hospital days (r = .39; p = .031), SGRQ (r = .36; p = .016), SCAQ (r = .41; p = .019), and 6-minute walking distance (r = .54; p = .030) were significantly different. The increase in the number of unscheduled outpatient visits in the intervention group during follow-up was attributed to acute exacerbations and a significant decrease in the number of hospitalizations and hospital days. Hence, the telenursing intervention program may be effective in reducing exacerbations in patients with chronic respiratory failure receiving NPPV at home.

**Trial Registration:** UMIN Clinical Trials Registry (UMIN-CTR) UMIN000027657

## Introduction

In recent years, as Japan’s aging population has been increasing more rapidly than any other country, there has been a call for the establishment of a comprehensive regional care system [1–3]. Japan is planning to shift approximately 300,000 hospital beds to home care by 2025 and is rapidly transitioning from hospital-based to community-based care [4-5].

A means to support the operation of a comprehensive community care system is telemedicine using information and communication technology (ICT) [6]. In Europe and the United States, telemedicine has become remarkably widespread and has been effective in reducing the cost of home health care by providing the same medical and nursing care to patients in their own residence [7]. Telemedicine includes telenursing, in which nurses provide health consultations with patients [8-10] and aims to improve the health of patients. Telenursing collects biological information and provides opportunities for accurate health consultation and guidance [11]. Telenursing is gradually spreading in Japan. The International Council of Nurses defines telenursing as the use of telecommunications technology in nursing to enhance patient care [12]. Thus far, Western countries have provided telenursing to patients with chronic obstructive pulmonary disease (COPD) and chronic heart failure, and telenursing has been effective in reducing the number of emergency visits, the length of hospital stay, and the number of acute exacerbations [13-17]. In Japan, telemedicine is provided to patients with COPD who are receiving home oxygen therapy, patients with diabetes, and patients with cancer. It can improve cost-effectiveness, the incidence of acute exacerbations, and quality of life (QOL) [8-10].

Chronic respiratory failure is a condition in which respiratory failure persists for more than 1 month [18]. The most common form of chronic respiratory failure is COPD, which is predicted to become the third leading cause of death by 2030 [19]. Many COPD patients are receiving home oxygen therapy (HOT) and noninvasive positive pressure ventilation (NPPV) [20]. Because NPPV does not involve tracheal intubation, patients are able to talk, eat, and drink as they go about their daily lives. The provision of NPPV has significantly improved the life expectancy and QOL of patients [21]. However, patients with chronic respiratory failure receiving NPPV experience a reduced range of activity owing to respiratory symptoms and distress when wearing a mask; patients receiving NPPV are more prone to have high carbon dioxide levels and need to learn to manage their physical condition to a higher degree [22]. Thus, patients with chronic respiratory failure receiving NPPV are more likely to have acute exacerbations owing to poor lung function reserve and exercise tolerance [23]. Chronic respiratory failure patients are less likely to notice changes in acute exacerbations because they usually have respiratory symptoms. This means that they delay seeking medical attention, which may lead to a more serious condition. Hospitalization due to acute exacerbation decreases activities of daily living (ADL) and contributes in lowering patient QOL [21]. Acute exacerbation in patients with chronic respiratory failure involves the sudden worsening of respiratory symptoms, such as dyspnea, cough, and sputum, and respiratory failure status [23]. According to the Japanese COPD guidelines, 50–70% of acute exacerbations are caused by respiratory infections, such as the common cold, 10% by air pollution, and 30% by unknown causes [24]. Harrison et al. reported that chronic respiratory failure patients can avoid acute exacerbations through effective self-management [25]. Self-management means that patients have the knowledge and skills to cope with their own symptoms and signs, while coping with their lives [26, 27]. A meta-analysis of COPD patients showed that self-management education is effective in reducing shortness of breath, improving health-related QOL, and decreasing the number of respiratory-related hospitalizations [28]. We hypothesized that nursing support for learning self-management in all aspects of convalescent life is necessary.

Telenursing for COPD patients receiving HOT by nurses was reported to be effective in preventing acute exacerbations and increasing physical activity [8]. However, the benefits of telemedicine nursing for chronic respiratory failure patients receiving NPPV, who are more dependent on medical care, has limited evidence. Therefore, this study will examine the effectiveness of a telenursing intervention program that combines non-pharmacological therapies such as symptom management and infection prevention behaviors for chronic respiratory failure patients receiving NPPV.

This study aimed to examine the effectiveness of a telenursing intervention program in reducing exacerbations in patients with chronic respiratory failure who are receiving NPPV at home using a randomized controlled trial. Our findings confirm our hypothesis that the proposed 3-month telenursing intervention program decreases the number of hospitalizations due to acute exacerbations, hospital days, and unscheduled outpatient visits in the intervention group.

## Materials and Methods

### Study design and setting

The study design was a randomized controlled trial with an intervention group using an ICT-based telenursing intervention program for chronic respiratory failure patients receiving NPPV at home in addition to their usual care and a control group providing usual care.

### Participants

The study participants included patients with chronic respiratory failure receiving NPPV attending respiratory outpatient clinic from September 2017 to September 2018. The inclusion criteria were as follows: chronic respiratory failure due to respiratory diseases; NPPV support; state of respiratory failure lasting more than one month, with a partial pressure of arterial blood carbon dioxide >45 mmHg; no specific disease and therefore cannot be staged; age ≥20 years; and ability to use a tablet device. The exclusion criteria were as follows: inability to communicate due to cognitive impairment; inability to speak Japanese; non-attendance at respiratory outpatient clinic.

### Treatment groups

The attending physician at the cooperating facility selected the subjects based on the eligibility criteria and explained the documents related to the study. An initial interview was conducted with participants who were interested in the study. After submitting a consent form for participation at the initial interview, participants were enrolled in the study. Consenting participants were assigned serial numbers, consolidated, and anonymized. Research assistants assigned the participants to the intervention and control groups by fitting them to a random number table generated from the serial numbers. Baseline data, including basic attributes and primary and secondary endpoints, were measured. The intervention group was given an overview of the telenursing intervention program and asked to input their physical condition, including vital signs, symptoms, and signs, using ICT at home. At the start of the program, we visited the participant’s home to check the operation of the tablet and set up the communication. Participants who consented to cooperate in the study were assigned serial numbers, consolidated, and anonymized. Research assistants assigned subjects to the intervention and control groups by fitting them to a random number table generated from the serial numbers. Those assigned to the intervention group were contacted by phone and given written and oral explanations of the intervention during home visits.

### Telenursing intervention program

The telenursing intervention program implemented in this study used a telenursing system (COMPAS) developed by the author. This program was also designed as a nursing intervention program to reduce acute exacerbations by integrating self-monitoring and self-management into the lives of chronic respiratory failure patients undergoing NPPV on a proactive basis according to their individual physical and mental conditions. The program consists mainly of telemonitoring and telehealth counseling using a telenursing system. The tablet device used by the participants were provided by the researcher. Once a day, each participant answered the questions displayed on the tablet from a list of options in approximately 10 min, and data on the physical condition, including vital signs, respiratory symptoms, food intake, excretion, medication use, physical symptoms other than respiratory symptoms, and questions to the medical staff, were sent to the server and collected. The transmitted data was monitored remotely, and nursing support was determined based on the results of each question item set in advanced consultation with the physician in charge.

When a change in physical condition was observed, the condition was assessed using videophone, telephone, or e-mail, and nursing support was provided based on the Respiratory Rehabilitation Manual [29]. The participants can input data into the tablet terminal from 08:00 to 12:00, and the nursing interventions were conducted as needed from 13:00 to 17:00 for approximately 30 minutes.

If there was failure to enter data on the tablet device for more than three days, the participant was contacted by phone to determine the cause. If the problem could not be solved, we immediately called back and addressed the issue as soon as possible.

Since the tablet device was provided by the researcher, they were collected following the completion of the intervention program, and all communication costs for this study were borne by the researcher. This program was supervised by a number of professionals during its development, including respiratory medicine specialists, nurses engaged in respiratory medicine, physical therapists, and occupational therapists.

### Telenursing system

This system was developed based on the researcher’s preliminary investigation [22] and literature review, and the web-based program comprised a patient site and a researcher site. The tablet of the patient site contains “daily physical status record,” “video phone (Skype),” and “respiratory rehabilitation information (PDF, YouTube videos)”. The researcher’s site included a list of the participants’ telemonitoring data and the ability to enter comments. The researcher could review the daily records of the participants and record the changes over time on a summary sheet (S1 Appendix. and S2 Appendix.).

The server was managed by a system production company and a system development company, which encrypted the transmitted data with Secure Socket’s Layer encryption. The researcher and the physician in charge of the cooperating medical institution were the only ones authorized to access the server. The communication technology used was Japanese cell phone communication. Near Field Communication (NFC) function was used for the collection of vital sign data. NFC is a short-range wireless communication technology that uses a frequency of 13.56 MHz and has a communication distance of approximately 10 cm. The NFC function assisted the subject to input data on vital signs.

### Survey method and contents

#### Characteristics of the participants

The following characteristics of the subjects were collected at the time of registration: age, sex, primary diagnosis, family composition, ADL, level of care required, implementation status of NPPV, implementation status of HOT, smoking history, and availability of social resources. The pulmonary ADL (P-ADL) was used as the ADL assessment scale. P-ADL was developed in Japan as a respiratory disease-specific ADL scale because the standard ADL scale does not accurately capture ADL impairment due to breathlessness in respiratory diseases [30].

#### Study endpoints

The primary endpoint of the study was the number of acute exacerbations in the past three months. For the number of acute exacerbations, the number of outpatient visits, hospital admissions, and hospital days were collected. The secondary endpoints included St. George’s Respiratory Questionnaire (SGRQ) score [31-33], Self-Care Agency Questionnaire (SCAQ) score [34-36], Euro QOL 5 Dimension (EQ-5D) score [37], pulmonary function tests, and six-minute walk distance (6MWD) [38]. These items were evaluated at enrollment and at follow-up (3 months after enrollment).

## Statistical analysis

The basic attributes of the subjects were examined for normality using the Kolmogorov– Smirnov test. Subsequently, Mann–Whitney’s U test and Pearson’s χ2 test were used to compare between the intervention and control groups. At the time of enrollment and follow-up, the endpoints of the intervention and control groups were compared using the Mann–Whitney’s U test. The number and content of interventions were tabulated. We used a personal computer with Microsoft Windows 8.1 operating system, and statistical analysis was performed using IBM SPSS Statistics ver. 21.0 software. Significance level was set at two-tailed p > 0.05

## Ethical considerations

The study protocol was approval of the Ethics Committee of the Tohoku University Graduate School of Medicine (approval number: 2017-1-400) and adhered to ethical principles of the “Declaration of Helsinki.” This study was conducted in compliance with the CONSORT guidelines. The trial registration number is UMIN-CTR (UMIN000027657). The study details, including an overview of the study objectives and other information, how to withdraw cooperation, protection of personal information, how to operate the tablet, encryption of transmitted data, and restrictions on access to the server, were adequately explained to the participants, and consent to participate was obtained orally and in writing.

The study provisions included the tablet and communication costs associated with the study, but not the electricity bill for using the tablet. Consolidated anonymization was used to protect participants’ personal information. The intervention program was conducted in a private room where privacy was maintained, and access to the room was restricted to non-researchers during data viewing, phone calls, and video calls. The researcher always logged out of the telenursing system when leaving the computer terminal.

## Results

### Participant inclusion

Of the 33 participants referred by the facilities, 31 (n=15, intervention group; n=16, control group) agreed to participate in the study. The two participants who did not agree to participate cited the following reasons: “too old to use a tablet device” and “unable to participate because of the survey schedule.” No participants dropped out after registration; thus, 31 participants were included in the analysis. The flowchart of participant inclusion is shown in Fig 1.

**Fig 1.**
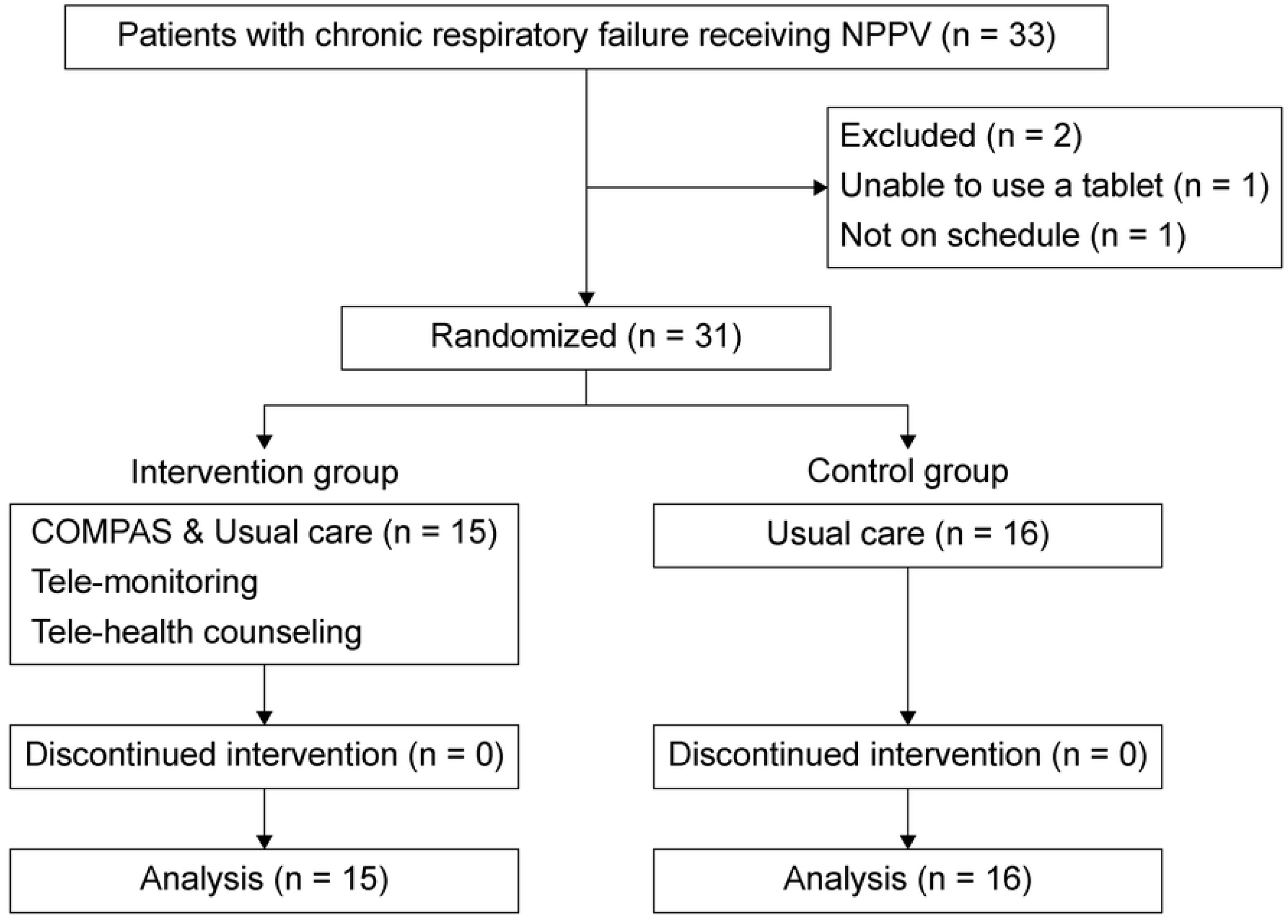
Flow diagram of participant inclusion

### Characteristics of the study participants

The characteristics of the subjects are shown in Table 1. There were nine men (60.0%) and six women (40.0%) in the intervention group and nine men (56.3%) and seven women (43.8%) in the control group. The mean age of all participants was 73.0 (standard deviation [SD]: 10.2) years; mean height, 155.8 (SD: 9.5) cm; mean weight, 60.5 (SD: 12.7) kg; and mean body mass index, 25.1 (SD: 5.6).

**Table 1.** Characteristics of the study participants (N = 31)

| Evaluation items | Total |  | Intervention group |  | Control group |  | *p* |
| --- | --- | --- | --- | --- | --- | --- | --- |
|  | *N*=31 |  | *n*=15 |  | *n*=16 |  |  |
| Gender, *n* (%) |  |  |  |  |  |  | .833 |
| Male | 18 | (58.1) | 9 | (60.0) | 9 | (56.3) |  |
| Female | 13 | (41.9) | 6 | (40.0) | 7 | (43.8) |  |
| Age (years), mean±SD | 73.0 | ±10.2 | 72.5 | ±10.9 | 73.6 | ±10.0 | .648 |
| Body data, mean±SD |  |  |  |  |  |  |  |
| Length (cm) | 155.8 | ±9.5 | 153.8 | ±0.1 | 157.4 | ±10.7 | .355 |
| Weight (kg) | 60.5 | ±12.7 | 60.9 | ±15.3 | 60.2 | ±10.6 | .861 |
| BMI | 25.0 | ±5.6 | 25.9 | ±7.1 | 24.4 | ±4.2 | .759 |
| Duration of oxygen therapy, mean±SD |  |  |  |  |  |  |  |
| NPPV (years) | 3.0 | ±3.7 | 2.9 | ±3.3 | 3.2 | ±4.1 | .835 |
| HOT (years) | 2.0 | ±1.6 | 1.7 | ±1.6 | 2.3 | ±1.6 | .489 |
| ADL, mean±SD |  |  |  |  |  |  |  |
| P-ADL (points) | 169.5 | ±22.92 | 167.1 | ±23.0 | 171.8 | ±23.3 | .678 |
| Primary disease, *n* (%) |  |  |  |  |  |  |  |
| COPD | 13 | (41.9) | 6 | (40.0) | 7 | (43.8) | .607 |
| Pulmonary cell hypoventilation syndrome | 10 | (32.3) | 5 | (33.3) | 5 | (31.3) |  |
| Pulmonary tuberculosis sequelae | 6 | (19.4) | 2 | (13.3) | 4 | (25.0) |  |
| Scoliosis | 1 | (3.2) | 1 | (6.7) |  |  |  |
| Spinal caries | 1 | (3.2) | 1 | (6.7) |  |  |  |
| NPPV treatment schedule, *n* |  |  |  |  |  |  |  |
| (%) |  |  |  |  |  |  |  |
| Nocturnal | 31 | (100.0) | 15 | (100.0) | 16 | (100.0) | - |
| HOT, *n* (%) | 16 | (51.6) | 8 | (53.3) | 8 | (50.0) | .853 |
| Current smoking, *n* (%) | 1 | (3.2) | 0 | (0.0) | 1 | (6.3) | .325 |
| Past smoking, *n* (%) | 19 | (61.3) | 10 | (66.7) | 9 | (56.3) | .552 |
| With Family, *n* (%) | 28 | (90.3) | 13 | (86.7) | 15 | (93.8) | .505 |
| Social resources, *n* (%) | 13 | (41.9) | 4 | (26.7) | 9 | (56.3) | .095 |

Age, Body data, Duration of oxygen therapy, ADL: Mann-Whitney U test
Gender, Primary disease, NPPV treatment schedule, HOT, Current smoking, Past smoking, Past consumption, With Family, Social resources: $\chi^2$ test

COPD was the main cause of chronic respiratory failure in 13 patients (41.9%), followed by alveolar hypoventilation syndrome in 10 (32.3%), sequelae of pulmonary tuberculosis in six (19.4%), and scoliosis and spinal caries in one each. All patients were prescribed NPPV only at night during bedtime. Approximately half (n = 16, 51.6%) of the patients were on HOT. There were 19 past smokers (61.3%) and 30 current non-smokers (96.8%). A total of 28 patients (90.3%) were living with family members, and 13 (41.9%) were using social resources. The mean use duration of NPPV and HOT was 3.0 (SD: 3.7) years and 2.0 (SD: 1.6) years, respectively. The P-ADL score was 169.5 (SD: 22.9) points. There were no significant differences in the characteristics of the participants between the intervention group and the control group at enrollment.

### Study endpoints at enrollment

The primary and secondary endpoints at enrollment were compared between the intervention and control groups and presented in Table 2.

**Table 2.**
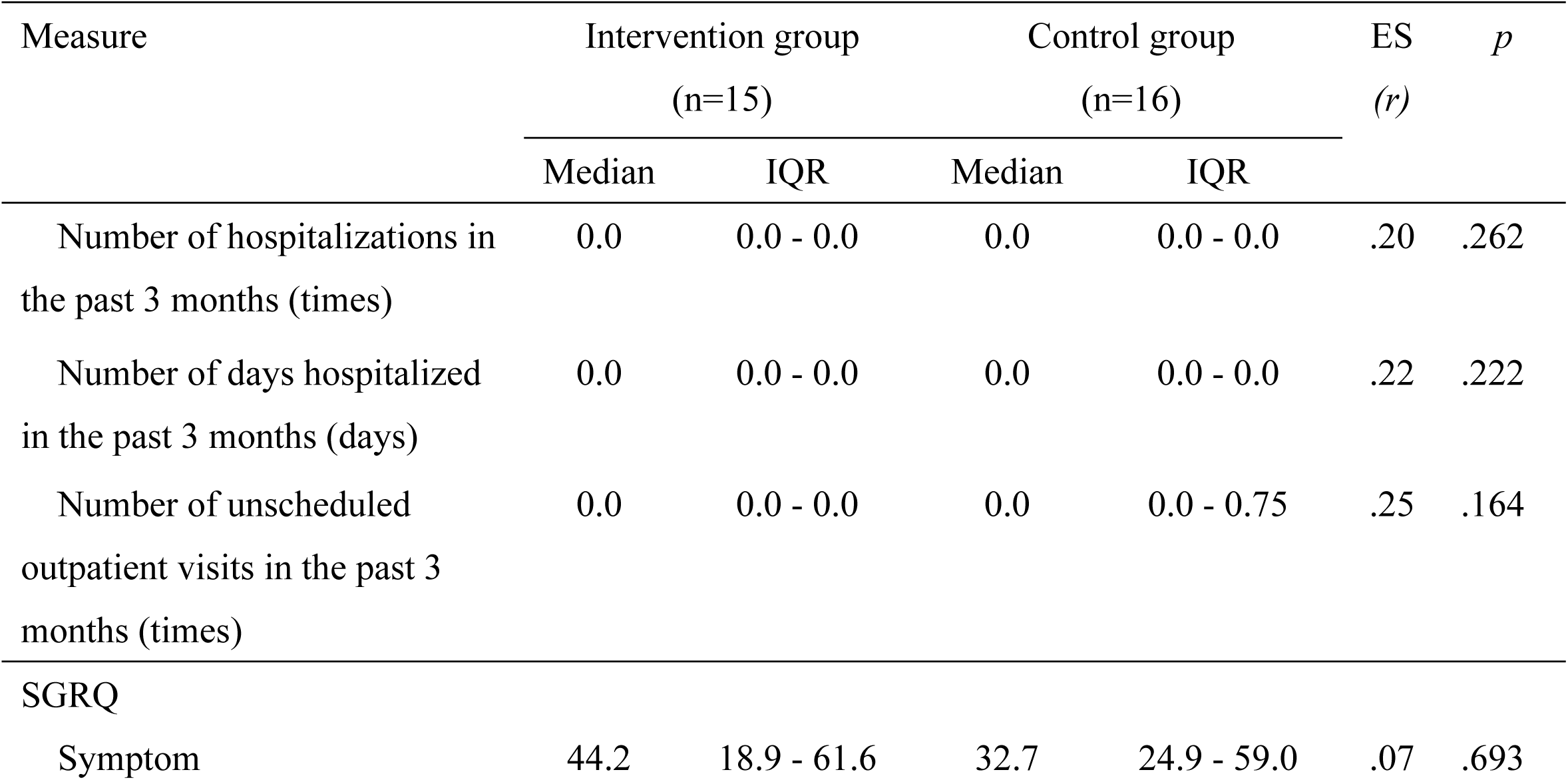
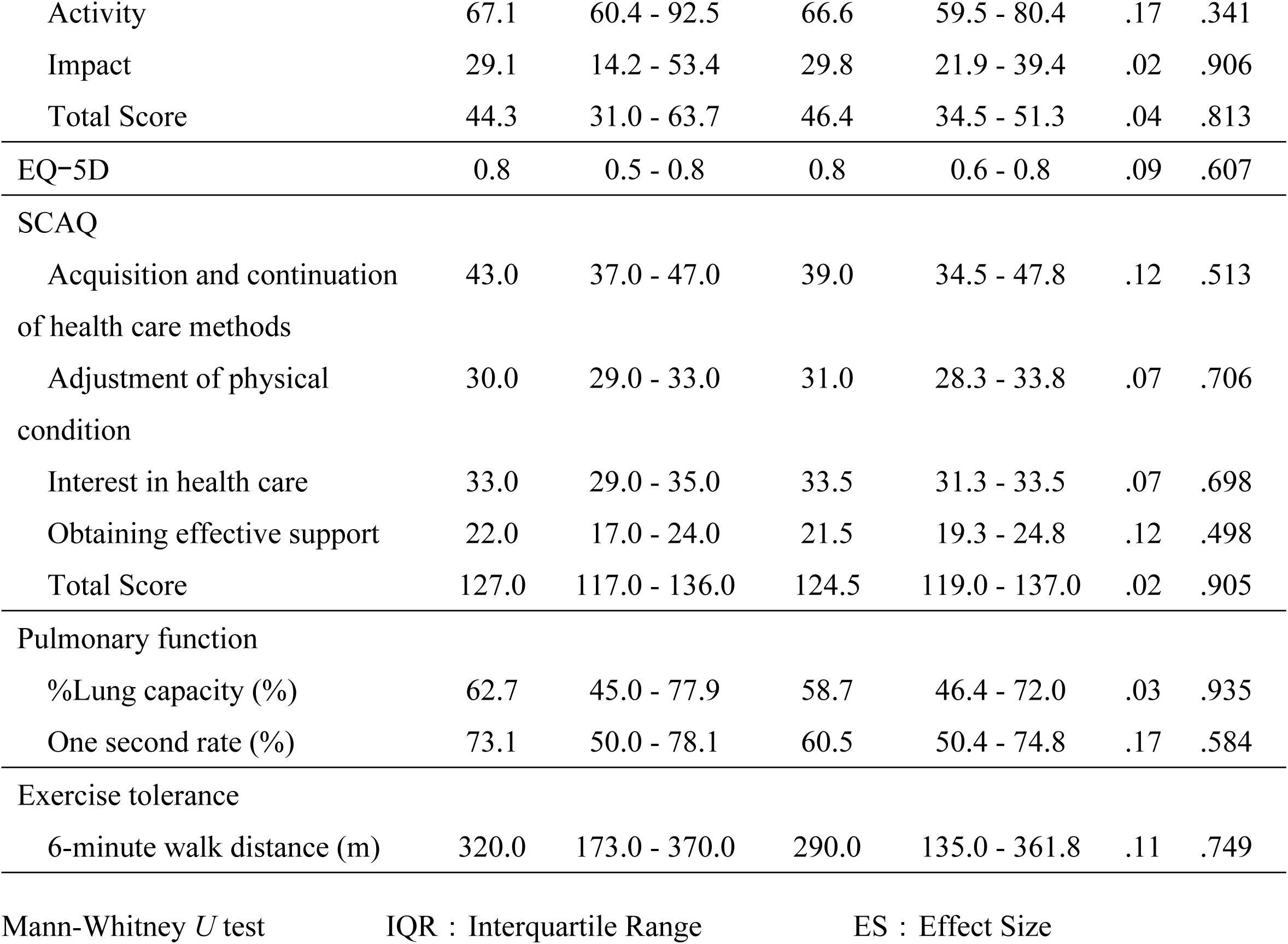
Assessment scores at enrollment.

There was no significant difference in the number of outpatient visits and hospitalizations and days of hospitalization between the intervention and control groups at enrollment.

Regarding secondary endpoints, the SGRQ score for the intervention group was 44.3 (IQR: 31.0–63.7) comprising a symptom score of 44.2 (IQR: 18.9–61.6), an activity score of 67.1 (IQR: 60.4–92.5), and an impact score of 29.1 (IQR: 14.2–53.4). The EQ-5D was 0.8 (IQR: 0.5–0.8), and the overall SCAQ was 127.0 (IQR: 117.0–136.0) comprising 43.0 (IQR: 37.0–47.0) for acquiring and maintaining health care practices, 30.0 (IQR: 29.0–33.0) for adjusting physical condition, 33.0 (IQR: 29.0–35.0) for interest in health care, and 22.0 (IQR: 17.0–24.0) for acquisition of effective support. The percent lung capacity of the pulmonary function test measurements was 62.7 (IQR: 45.0–77.9)%, the 1-second rate was 73.1 (IQR: 50.0–78.1)%, and the 6MWD was 320.0 (IQR: 173.0–370.0) m.

The control group has a total SGRQ score of 46.4 (IQR: 34.5–51.3), with a symptom score of 32.7 (IQR: 24.9–59.0), an activity score of 66.6 (IQR: 59.5–80.4), and an impact score of 29.8 (IQR: 21.9–39.4). The EQ-5D was 0.8 (IQR: 0.6–0.8), and the overall SCAQ score was 124.5 (IQR: 119.0–137.0), comprising 39.0 (IQR: 34.5–47.8) for acquisition and continuation of health care methods, 31.0 (IQR: 28.3–33.8) for adjustment of physical condition, 33.5 (IQR: 31.3–33.5) for interest in health care, and 21.5 (IQR: 19.3–24.8) for acquisition of effective support. The % lung capacity was 58.7 (IQR: 46.4–72.0)%, and the 1-second rate was 60.5 (IQR: 50.4–74.8)%. The 6MWD was 290.0 (IQR: 135.0–361.8) m. Overall, there was no significant difference in the secondary endpoints between the intervention and control groups at enrollment.

### Study endpoints at follow-up

The study endpoints at follow-up were compared and are shown in Table 3 and Fig 2. The intervention group had significantly fewer hospitalizations (r = .38; p = .037) and shortened hospital stay (r = .39; p = .031) than the control group. There was a significant increase in the number of unscheduled outpatient visits in the intervention group compared with the control group (r = 36; p = .045).

**Table 3.**
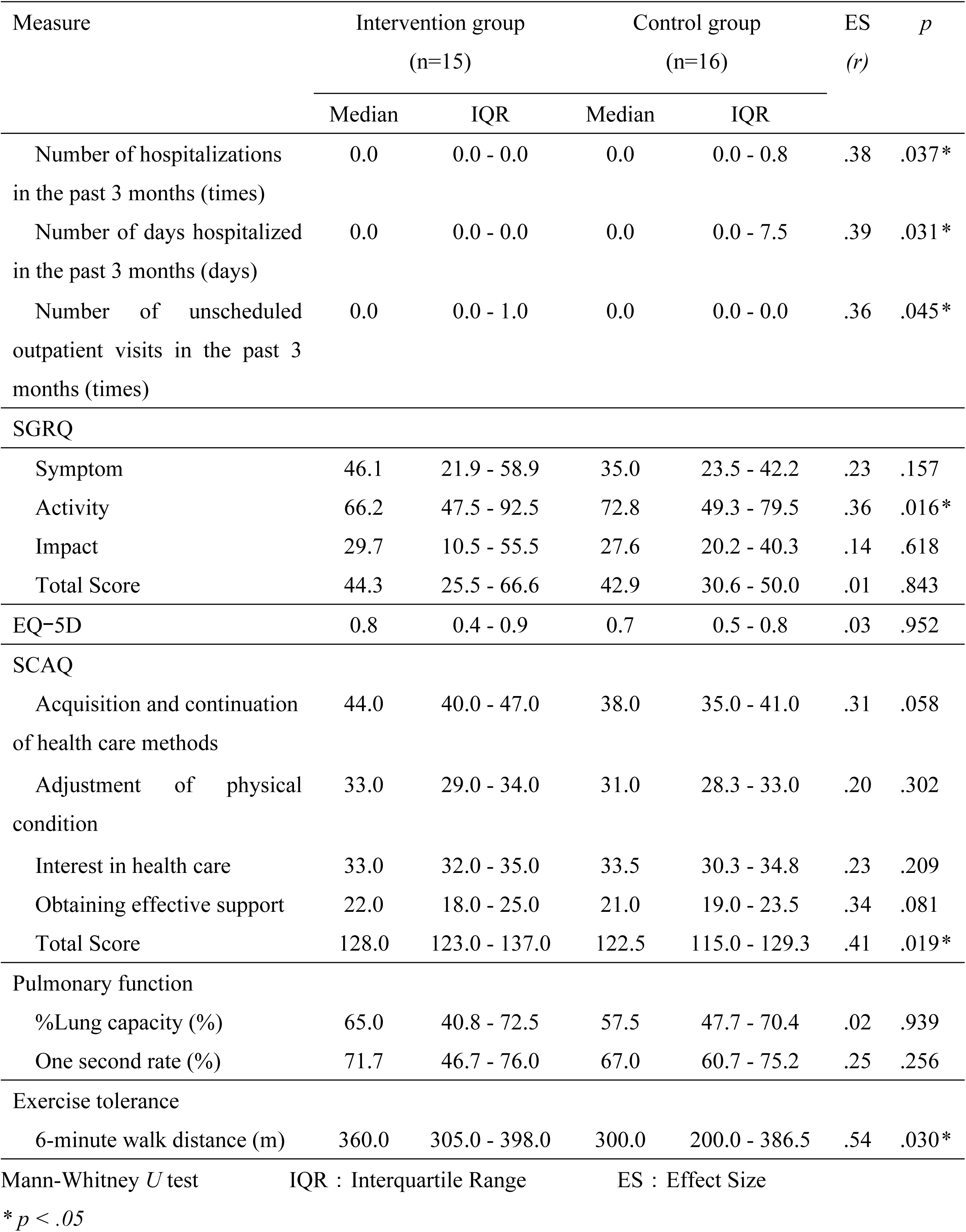
Assessment scores at three-month follow-up.

| Measure | Intervention group<br>(n=15) |  | Control group<br>(n=16) |  | ES<br>(*r*) | *p* |
| --- | --- | --- | --- | --- | --- | --- |
|  | Median | IQR | Median | IQR |  |  |
| Number of hospitalizations<br>in the past 3 months (times) | 0.0 | 0.0 - 0.0 | 0.0 | 0.0 - 0.8 | .38 | .037* |
| Number of days hospitalized<br>in the past 3 months (days) | 0.0 | 0.0 - 0.0 | 0.0 | 0.0 - 7.5 | .39 | .031* |
| Number of unscheduled<br>outpatient visits in the past 3<br>months (times) | 0.0 | 0.0 - 1.0 | 0.0 | 0.0 - 0.0 | .36 | .045* |
| SGRQ |  |  |  |  |  |  |
| Symptom | 46.1 | 21.9 - 58.9 | 35.0 | 23.5 - 42.2 | .23 | .157 |
| Activity | 66.2 | 47.5 - 92.5 | 72.8 | 49.3 - 79.5 | .36 | .016* |
| Impact | 29.7 | 10.5 - 55.5 | 27.6 | 20.2 - 40.3 | .14 | .618 |
| Total Score | 44.3 | 25.5 - 66.6 | 42.9 | 30.6 - 50.0 | .01 | .843 |
| EQ-5D | 0.8 | 0.4 - 0.9 | 0.7 | 0.5 - 0.8 | .03 | .952 |
| SCAQ |  |  |  |  |  |  |
| Acquisition and continuation<br>of health care methods | 44.0 | 40.0 - 47.0 | 38.0 | 35.0 - 41.0 | .31 | .058 |
| Adjustment of physical<br>condition | 33.0 | 29.0 - 34.0 | 31.0 | 28.3 - 33.0 | .20 | .302 |
| Interest in health care | 33.0 | 32.0 - 35.0 | 33.5 | 30.3 - 34.8 | .23 | .209 |
| Obtaining effective support | 22.0 | 18.0 - 25.0 | 21.0 | 19.0 - 23.5 | .34 | .081 |
| Total Score | 128.0 | 123.0 - 137.0 | 122.5 | 115.0 - 129.3 | .41 | .019* |
| Pulmonary function |  |  |  |  |  |  |
| %Lung capacity (%) | 65.0 | 40.8 - 72.5 | 57.5 | 47.7 - 70.4 | .02 | .939 |
| One second rate (%) | 71.7 | 46.7 - 76.0 | 67.0 | 60.7 - 75.2 | .25 | .256 |
| Exercise tolerance |  |  |  |  |  |  |
| 6-minute walk distance (m) | 360.0 | 305.0 - 398.0 | 300.0 | 200.0 - 386.5 | .54 | .030* |

Mann-Whitney *U* test IQR : Interquartile Range ES : Effect Size
\* *p* < .05

**Fig 2.**
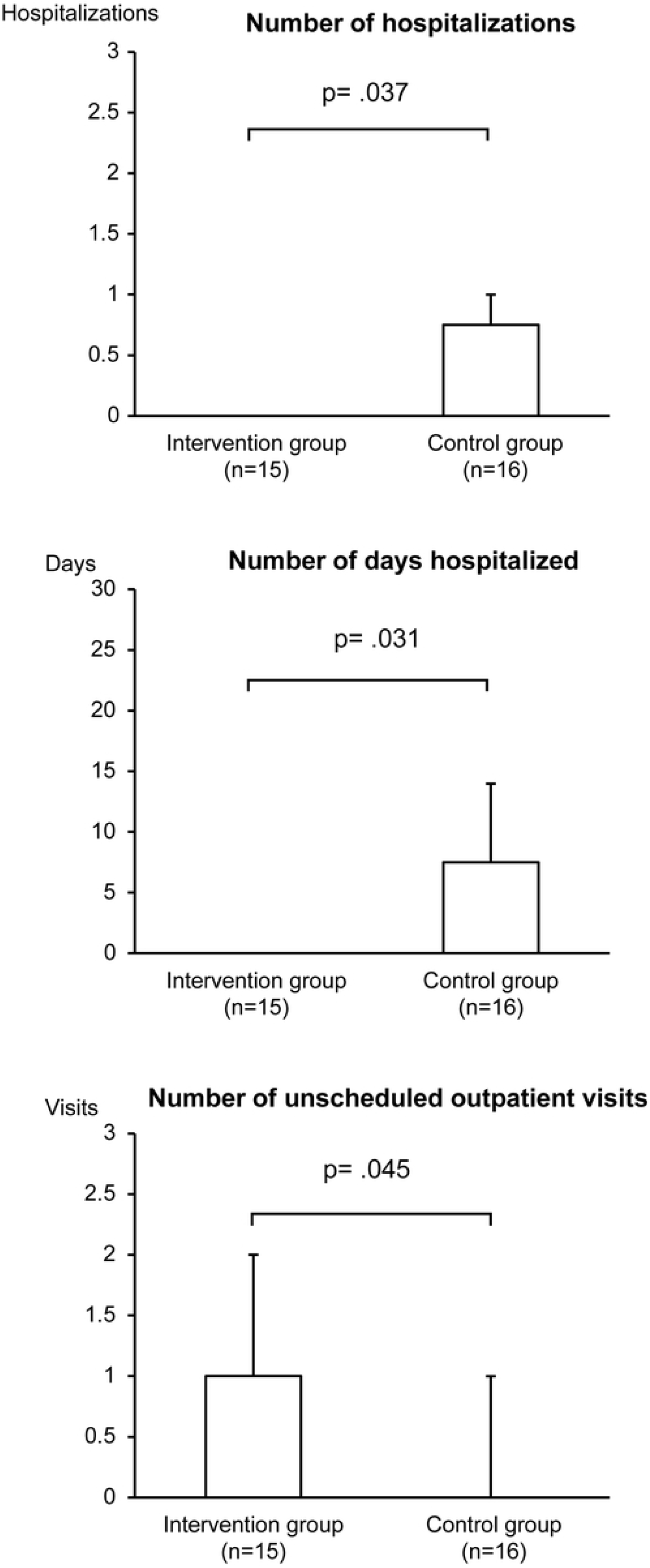
Primary endpoints at three-month follow-up

Group comparisons of the differences in secondary endpoints revealed a significantly lower SGRQ “activity” score in the intervention group than in the control group (r = .36; p = .016). This indicates that the intervention group had a higher QOL regarding activity. The SCAQ total scores of the intervention group were significantly higher than those of the control group (r = .41; p = .019), indicating a higher capacity for self-care. The 6MWD of the intervention group was significantly higher than that of the control group (r = .54; p = .030), indicating a higher exercise tolerance.

### Telehealth counseling

The total number of telehealth counseling sessions throughout the study period was 35, with an average of 2.3 consultations per participant (Table 4). There were 13 videophone consultations, 12 telephone consultations, and 10 e-mail consultations. There were 16 telehealth counseling sessions via remote monitoring. The most common reason for consultation was deterioration of vital signs (n = 6). The number of interventions based on the patient’s request was 19. There were six consultations regarding physical activity and four consultations regarding dyspnea.

**Table 4.** Data on telehealth counseling.

|  |  |
| --- | --- |
| Number of health consultations and information provided via videophone, etc. |  |
| Total | 35 |
| Average number of cases per person | 2.3 |
| Details |  |
| 1) Videophone | 13 |
| 2) Phone | 12 |
| 3) Email | 10 |
| Contents of health consultation and information provided by videophone, etc. |  |
| 1. telemonitoring intervention |  |
| Total | 16 |
| Details |  |
| 1) Deterioration of vital signs | 6 |
| 2) Deterioration of non-respiratory symptoms | 4 |
| 3) Deterioration of respiratory symptoms | 1 |
| 4) Delayed input | 5 |
| 2. intervention at the patient's request |  |
| Total | 19 |
| Details |  |
| 1) Physical activity | 6 |
| 2) Dyspnea | 4 |
| 3) NPPV device or mask | 3 |
| 4) Eating | 2 |
| 5) Other | 4 |

## Discussion

This study examined the effectiveness of a telenursing intervention program in reducing exacerbations in chronic respiratory failure patients receiving NPPV using a randomized controlled trial. The telenursing intervention program used in this study consisted of telemonitoring and telehealth counseling.

The number of hospitalizations for acute exacerbations and the duration of hospital stay at 3-month follow-up were reduced in the intervention group, suggesting that the proposed telenursing intervention program is effective in reducing acute exacerbations in chronic respiratory failure patients receiving NPPV. Previous studies using ICT have similarly reported a reduction in the number of hospitalizations due to acute exacerbations in patients with chronic respiratory failure [8, 13, 39]. Since there are no reports when limited to chronic respiratory failure patients receiving NPPV, the results of this study could be an important finding.

### Effectiveness of telemonitoring

We found that chronic respiratory failure patients receiving NPPV routinely experience respiratory symptoms, symptoms of hypercapnea, and pressure of the face mask during their recuperation. Some participants recognized the various symptoms as age-related changes or those of the common cold and took necessary coping actions, such as resting and monitoring of symptoms. Some participants thought that they should not attempt outpatient visits outside of their regular appointments and chose to wait until the next regular outpatient visit, despite having symptoms of an acute exacerbation. These results indicate that although signs of an acute exacerbation were detected, participants did not take prompt medical attention. The SCAQ, which indicates self-care ability, was significantly higher in the intervention group than in the control group, suggesting that the intervention program may have improved the self-management ability of the participants.

When the participants experienced a change in physical condition, they were instructed to contact the researcher for a telehealth counseling session. The researcher confirmed the results of the telemonitoring and explained the meaning of the symptoms and coping behaviors, which may have led the participant to learn the meaning of the change in physical condition and visit an outpatient clinic. According to previous studies using ICT, the number of visits to the emergency room was reduced because patients were able to receive direct instructions on medical care to be provided at home from medical personnel as a result of telemonitoring [40–42]. In a previous study, the participant had type II chronic respiratory failure, which is a complex condition that requires immediate and accurate medical intervention [24]; thus, the attending physician instructed the participant to seek immediate medical attention if any change in physical condition indicating an acute exacerbation was observed. Signs of an acute exacerbation include not only the appearance of clear respiratory symptoms but also sensations that are somehow different from the norm [20, 43].

It is difficult to link these subtle sensations to therapeutic actions, and many participants decided to wait and observe symptom progression without consulting a doctor. Symptoms of acute exacerbations, such as shortness of breath, sputum volume, and changes in sputum coloration, rarely appear more than once. Because they are short-lived, they are not easily judged to be acute exacerbations [24]. However, early detection and treatment of signs of acute exacerbations are important to prevent progression to acute exacerbations that require hospitalization. A previous study on showed that the number of nurse visits in the telemonitoring group increased, although the hospitalization rate of patients decreased [44]. Telemonitoring is considered to be important not only for early detection of acute exacerbations, but also for connecting patients to medical intervention at an early stage and reducing exacerbations.

Patients with type II respiratory failure are treated with pharmacotherapy and non-pharmacological therapies, such as respiratory rehabilitation from the stable stage. The emergence of acute exacerbations is related to complex pathological conditions, such as CO_2_ narcosis and pulmonary heart disease, and delay in detection of the signs of acute exacerbations and failure to respond to them directly led to deterioration of prognosis [21, 24]. Nurses who provide telenursing to patients with type II respiratory failure should monitor the results of telemonitoring, collaborate with the attending physician, and carefully consider the method of medical intervention.

### Effectiveness of telehealth counseling

Generally, NPPV patients receive self-management support from medical staff on how to manage NPPV equipment and knowledge and skills on medical treatment. They are then discharged home. However, in recent years, there are many elderly patients with cognitive decline in the latter half of their lives, and there are cases where it is difficult for them to utilize the knowledge and skills they have received at home. In this program, participants entered information about NPPV management, medication, diet, excretion, and physical activity, and the researcher provided telehealth counseling for questions and concerns about daily life. As a result, the participants were able to reconfirm the points to keep in mind during their medical treatment and may have been able to maintain their health management behavior.

Patients with respiratory diseases who continue appropriate self-management have reduced shortness of breath, improved health-related QOL, and decreased number of respiratory-related hospitalizations [45]. Since telenursing is conducted in a face-to-face format via videoconference, it is easier for patients to feel at ease and build a trusting relationship [46]. Telehealth counseling imparted the participants a strong sense of connection with the researcher. In addition, because it was easy to understand the living conditions of the participants, it was possible to assess their self-care abilities and provide tailored information although they were living far away.

Patients living with NPPV at home face challenges and questions that they did not face during their hospitalization. Telehealth counseling allowed participants to respond immediately to any changes in symptoms or questions about treatment. As a result, participants were able to cope with the situation with ease and acquire new knowledge and skills from the experience. For effective self-management support, behavioral change theory is used, and it is important to provide education and explanation in a timely manner when the patient is conscious [29, 47]. In this program, the timing of the telehealth counseling was not decided in advance, and it is possible that the program might have promoted smoother behavioral change in patients by providing counseling when they needed it.

Self-management education using ICT has been practiced in Japan and abroad and has been reported to be effective in reducing the number of hospitalizations and maintaining adherence [48, 49]. In this study, the intervention group had higher SCAQ scores and showed better maintenance of self-care capability than the control group. In other words, it is inferred that new knowledge and skills were added by the self-management support at the appropriate time. In addition, acute exacerbation may have been reduced by the continuation of appropriate health management strategies, such as in terms of food, excretion, and medication, during treatment.

### Future development of telenursing

The causes of acute exacerbations in patients with chronic respiratory failure mainly include respiratory infections and environmental factors; however, in approximately 30% of cases, the cause remains unknown [24]. Patient-reported exacerbations were the most frequently reported, and it is important for patients to learn how to assess and cope with acute exacerbations by themselves [50]. We demonstrated that a telenursing intervention program consisting of telemonitoring and telehealth counseling sessions reduced the number of hospitalizations and duration of hospital stay. Thus, we believe that telenursing is an effective way to help patients recognize the signs of acute exacerbations and to impart coping strategies at an early stage.

To effectively prevent the severity of acute exacerbations in patients with chronic respiratory failure, it is necessary to further improve the quality of regular medical care and outpatient nursing care and to promote telenursing. In recent years, setting up an action plan according to the patient’s symptoms has been considered effective to prevent acute exacerbations of COPD [51-53]. In particular, when patients become aware of signs of an acute exacerbation, they should take antimicrobial agents or oral steroids as instructed by their physicians [51]. More than half of the participants in this study did not have an action plan, and the physician instructed the patient to attend an outpatient visit promptly at the first sign of an acute exacerbation. However, there were times when the participant avoided visiting the outpatient clinic because of the burden of travel expenses, time, and physical exhaustion. Thus, it is necessary to integrate existing medical care and telenursing systems to complement each other, so that the burden on patients can be reduced and the severity of acute exacerbations can be prevented.

### Limitations of the study

This study had some limitations. First, since the participants were mainly older adults aged ≥60 years, it took some time to learn how to use the tablet devices. In addition, although the telenursing system used in this program was created for older adults, several home visits were required because the participants could not address by themselves issues regarding the communication environment or the applications. This may have had some impact on the evaluation of this study. Future studies should improve the communication environment and the methods of the system to make it more convenient for elderly patients. In comparison with a previous study [51], the follow-up period in this study (3 months) may have been too short for the participants to make behavioral changes in their lifestyle [47]. Thus, the longer-term effects of this intervention should be further investigated.

## Conclusion

The effectiveness of a telemedicine intervention program to reduce exacerbations in chronic respiratory failure patients receiving NPPV was evaluated in this randomized controlled trial with 15 patients in the intervention group and 16 patients in the control group. After three months of exposure to the telenursing intervention program, both the number of hospitalizations for acute exacerbations and the number of hospital days decreased. In addition, there was an improvement in the self-care ability of the patients in the intervention group. Therefore, the telemedicine intervention program may reduce exacerbations in chronic respiratory failure patients receiving NPPV.

## Data Availability

All relevant data are within the manuscript and its Supporting Information files.

## Acknowledgments

We would like to express our sincere gratitude to all the participants and their families who cooperated in this survey. We would also like to express our gratitude to the doctors, nurses, and other co-medical staff at the facilities that cooperated in the survey.

## Supporting Information

**S1 Appendix. Overview of the telenursing system (COMPAS)**

**S2 Appendix. Screen display of the telenursing system (COMPAS)**

